# STHLM3-F/U Study Protocol: Long-term effects on prostate cancer mortality after the Stockholm3 prostate cancer screening trial (STHLM3)

**DOI:** 10.1101/2025.02.13.25320514

**Authors:** Chiara Micoli, Andrea Discacciati, Martin Eklund

**Affiliations:** Department of Medical Epidemiology and Biostatistics, Karolinska Institutet, Stockholm, Sweden

## Abstract

**Background:** Prostate cancer (PCa) is the leading cause of cancer death among men in Sweden. Although PSA (prostate-specific antigen) testing to screen for prostate cancer is common, its use remains controversial. This debate is primarily fueled by its insufficient operating characteristics, resulting in many benign biopsies and overdiagnosis of indolent disease. The population-based diagnostic STHLM3 trial developed and validated the Stockholm3 test, demonstrating that the combined use of PSA and Stockholm3 can identify clinically significant PCa with higher specificity while retaining the same sensitivity as PSA alone. However, the long-term implications of screening with the combined use of PSA and Stockholm3 on mortality remain unknown.

**Aim:** This study aims to evaluate the effect of a one-time *invitation* for prostate cancer screening (using PSA and Stockholm3 test in combination) in men aged 50–69 living in Stockholm on prostate cancer mortality and all-cause mortality, as well as its effects on prostate cancer incidence, PSA testing, and biopsy rates. The research question investigated in this study (STHLM3-F/U) concerns one of the pre-specified secondary outcomes of the original population-based diagnostic STHLM3 study (ISRCTN84445406).

**Randomization and Intervention:** The trial involves men aged 50 to 69 living in Stockholm at the moment of invitation and who did not have a prostate cancer diagnosis. Randomization occurred at the population level using registry-based information, eligible men were randomly assigned to receive an invitation for screening or to continue with standard care. Invited men who decided to participate in the STHLM3 trial took a PSA test, followed by the Stockholm3 test if PSA level was above 1 ng/mL. Systematic biopsy was recommended for those with PSA ≥ 3 ng/mL and/or a Stockholm3 score ≥ 10%. The non-invited group did not receive any invitations and relied on standard care.

**Outcomes and analysis:** Primary outcome was prostate cancer mortality. Secondary outcomes included all-cause mortality, prostate cancer incidence, PSA testing, and biopsy rates. Data will be analyzed using registry information, with primary analysis using follow-up data until 2023-12-31. Time to prostate cancer mortality will be analyzed using the Aalen-Johansen estimator. Cumulative risks at the end of the follow-up will be compared between the invited and non-invited groups by means of a Risk Ratio at an alpha level of 3.5%. Both intention-to-screen and compliance-adjusted analyses will be performed.

**Discussion:** This study aims to address the knowledge gap regarding the long-term effect of screening with PSA and Stockholm3 tests in combination and provide insights into its potential effects on preventing mortality. Understanding whether a systematic organized screening intervention, using a combination of PSA and Stockholm3 has any effect on mortality can help inform decisions on prostate cancer screening policies.

**Ethics and dissemination:** Approval for ethical conduct has been obtained from the Stockholm regional ethics committee. Results will be shared through peer-reviewed publications.

## 1. Introduction

This STHLM3-F/U protocol describes the background and planned analyses for the long-term follow-up of the STHLM3 prostate cancer diagnostic trial (ISRCTN84445406)^1^.

### 1.1 Background and Rationale

Prostate cancer is a common disease, affecting approximately 1 in 8 men in the Western world during their lifetime^2^. In Sweden, prostate cancer is the primary cause of cancer mortality in men, with more than 2000 deaths every year^3^. The disease imposes a substantial burden on the healthcare system, with more than 120,000 men in Sweden living with a prostate cancer diagnosis. While low-risk prostate cancers are associated with low mortality rates, more aggressive forms impose a marked risk of cancer mortality and account for the majority of prostate cancer related deaths. Screening aims to reduce mortality by identifying cancers at an early stage, before they develop to a point when curative treatment is no longer possible. For prostate cancer, screening has over the last 30 years primarily relied on the use of the prostate-specific antigen (PSA) test^4^. However, prostate cancer screening using PSA is controversial, due to high rates of detection of low-risk prostate cancer typically associated with overdiagnosis, as well as high rates of unnecessary biopsies (benign findings)^5^. Different diagnostic tests are available that could mitigate PSA limitations, like the Stockholm3 test. Recently, magnetic resonance imaging (MRI) has also been introduced to reduce diagnosis of clinically low-risk prostate cancer (typically regarded as overdiagnosis) and the number of men who undergo biopsy unnecessarily.

### 1.2 The STHLM3 trial

The population-based STHLM3 prostate cancer diagnostic study^6^ (ISRCTN84445406) was conducted from May 2012 to January 2015 (the data lock for the primary analysis of the trial was done in December 2014^6^; some invited participants joined the trial after this data lock and were included in a subsequent analysis^7^), and was a one-time intervention that used a combination of a PSA and a Stockholm3 test to screen for clinically significant prostate cancer (defined Gleason Score ≥7 using 10-12 core systematic biopsy). The Stockholm3 test is a prediction algorithm that uses clinical variables, plasma protein biomarkers, and a polygenic risk score^6,8^ (Figure 1). The STHLM3 trial showed that the use of the Stockholm3 test, compared to PSA alone, can identify clinically significant prostate cancer with higher specificity while retaining the same sensitivity. Furthermore, the study showed that using the Stockholm3 model could reduce the number of men diagnosed with a Gleason Score 6 cancer and the number of men needing to undergo biopsy. The use of the Stockholm3 test serves as a more accurate method to find prevalent prostate cancers in a population of asymptomatic men. It should be noted that opportunistic PSA based screening was common in the population of men aged 50-69 years old in Stockholm at the time of the STHLM3 trial, with about one third of the men in the population (and two thirds among STHLM3 participants) having undergone at least one previous PSA test.

**Figure 1.**
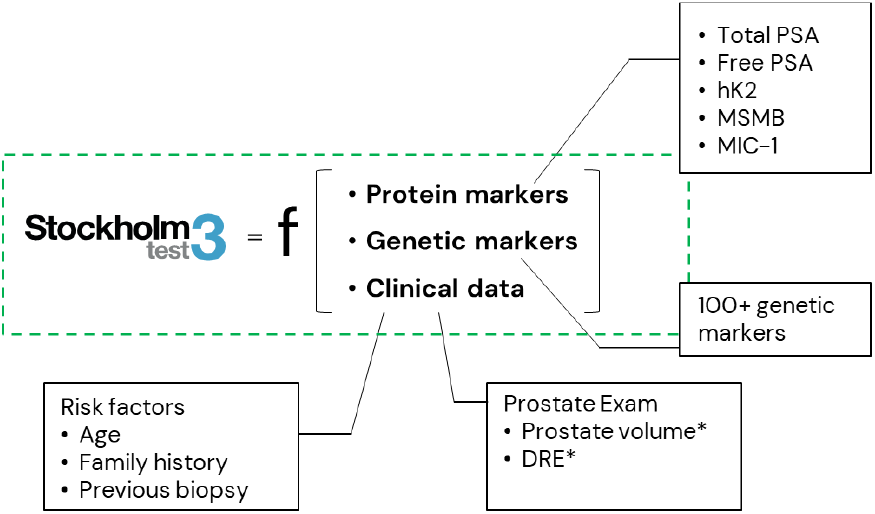
Conformation of the Stockholm3 test developed and used in the STHLM3 trial.

In the STHLM3 trial, eligible men (age 50 to 69 and residing in the Stockholm area with a permanent postal address, without previous prostate cancer diagnosis) were randomly assigned to either receive an invitation to undergo a single prostate cancer test (i.e. a combination of PSA and Stockholm3) or to not be invited and to continue with usual standard of care (Figure 2).

**Figure 2.**
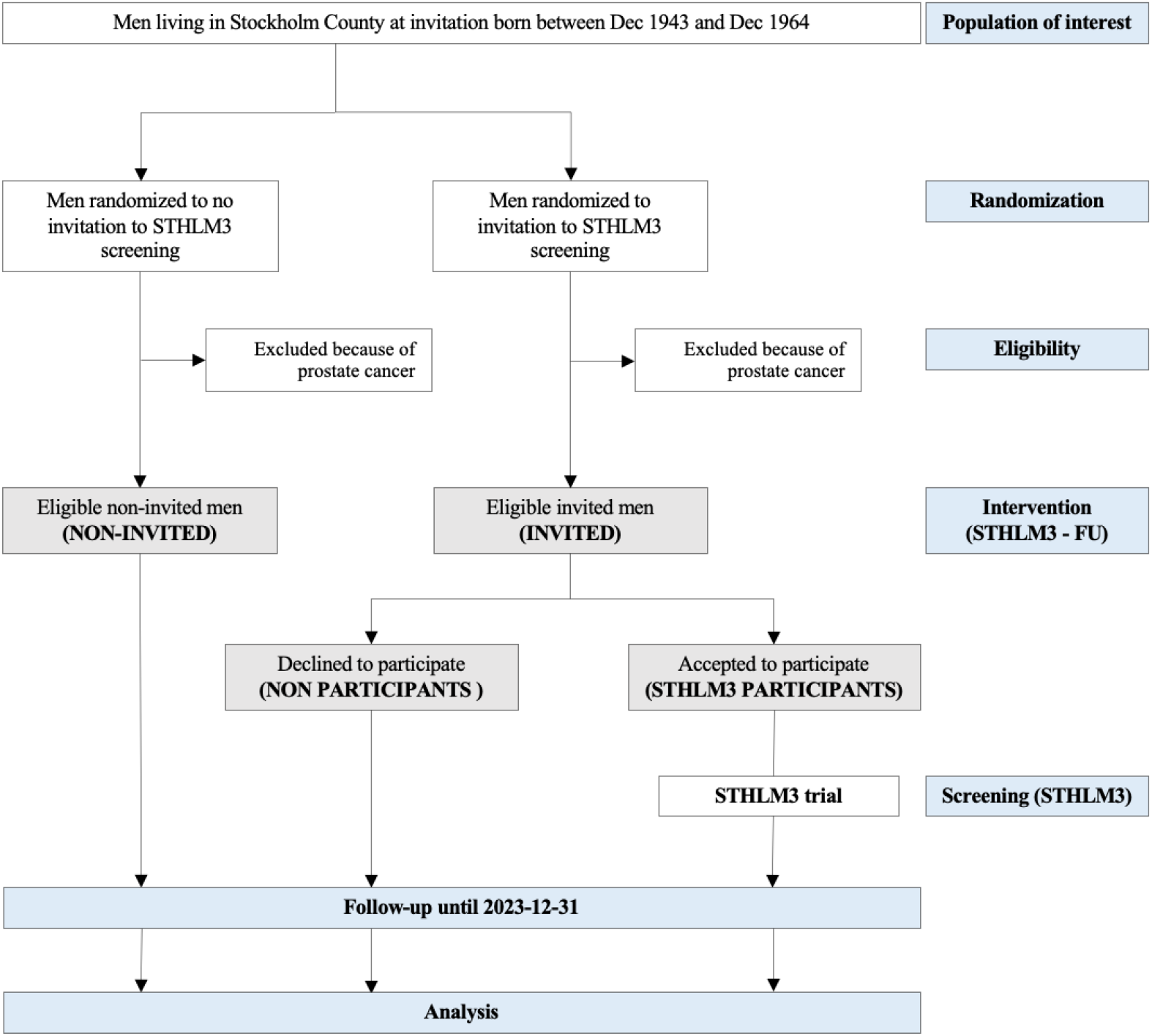
CONSORT diagram

From the pool of eligible candidates, a random selection was made from the Swedish Population Register maintained by the Swedish Tax Agency, and these individuals were contacted via mail with an invitation to participate in the STHLM3 trial.

For the invited men who responded to the invitation, the STHLM3 screening intervention involved two sequential steps: initially, all participants underwent a PSA test. Those with a PSA value ≥1 ng/mL then underwent the Stockholm3 test. Men with a PSA level ≥3 ng/mL or a Stockholm3 value ≥10% (or both) were recommended to undergo systematic prostate biopsy (STHLM3 was conducted before the introduction of pre-biopsy MRI and targeted biopsy in the Stockholm Region).

Following negative PSA/Stockholm3 tests (i.e. a PSA <3 ng/mL and a Stockholm3 test <10%), a recommendation of when to return for testing was sent to study participants by mail. The recommendation was based on individual test values (re-test in 6-10 years if PSA <1 ng/mL, retest in 2 years if PSA ≥1 ng/mL and PSA <3 ng/mL and Stockholm3 test <10%). Everything else (including treatment if diagnosed) was left to standard follow-up and treatment management was conducted as standard of care through the healthcare system.

The comparison group did not receive any type of invitation and continued undergoing the usual standard of care (Figure 2). Standard of care in Stockholm involved prevalent opportunistic testing, with about 25% of the population aged 50-59 and 40% of aged 60-69 having undergone at least one PSA test in the two years before the STHLM3 trial^9^ (and 66% of STHLM3 participants having undergone at least one previous PSA test before enrolling in the STHLM3 trial).

Thus, the STHLM3 study constituted of two nested interventions: (1) One randomized intervention where men were randomly assigned to receive by mail an invitation to undergo prostate cancer screening using the PSA and Stockholm3 test; and (2) One intervention using a paired screen-positive design where the test characteristics of the two tests (PSA and Stockholm3) were compared in men who were randomized to receive the STHLM3 screening invitation *and* who decided to participate (main analysis for this intervention was published in the Lancet Oncology in 2015^6^). This protocol describes how we will investigate the long-term effects of the first intervention, i.e. the one-off invitation to prostate cancer screening using both PSA and Stockholm3 (STHLM3-F/U).

## 2. Methods

### 2.1 STHLM3-F/U research questions

This study aims to evaluate the effect of a one-time *invitation* for prostate cancer screening (using PSA and Stockholm3 in combination) in men aged 50–69 living in Stockholm on prostate cancer-specific and all-cause mortality, as well as its effects on prostate cancer incidence, PSA testing, and biopsy rates.

The primary outcome of this study is prostate cancer mortality. Secondary outcomes will include all-cause mortality, prostate cancer incidence, distinguishing between subgroups based on Gleason Score at diagnosis. PSA and biopsy patterns in the two randomized groups will also be evaluated.

All endpoints will be collected using registry data^9^. The primary analysis will be conducted with follow-up data until 2023-12-31, with the possibility of repeating the analysis in the future (at 15 and 20 years of follow-up). More details about the follow-up are explained in Section 2.2.

#### STHLM3-F/U

- **Population (P):** Men aged 50 to 69 living in Stockholm county, without a prior prostate cancer diagnosis.
- **Intervention (I):** A single invitation to screen for prostate cancer utilizing PSA and the Stockholm3 test in combination. If a man tested positive on either PSA ≥ 3 ng/mL or Stockholm3 ≥ 10%, he was recommended to undergo standard systematic prostate biopsy.
- **Comparison (C):** Unscreened standard care, where ‘unscreened’ means that men in the comparison group were not invited to the STHLM3 trial (a single screen for prostate cancer using PSA combined with Stockholm3). Many men in the control group however undergo opportunistic prostate cancer screening (according to standard of care at the time of the STHLM3 trial and during the follow-up time).
- **Outcome (O):**
  - Primary outcome: Prostate cancer mortality.
  - Secondary outcomes: All-cause mortality, prostate cancer incidence, PSA and biopsy patterns.
- **Timeframe (T):** Analysis conducted with follow-up until 2023-12-31, with the possibility of repeating the analysis at 15 and 20 years of follow-up.

### 2.2 Randomization and follow-up

The research question investigated in this study (STHLM3-F/U) concerns one of the original secondary outcomes of the population-based diagnostic STHLM3 study, prespecified in the original statistical analysis plan^6^.

For the STHLM3-F/U intervention, randomization was conducted at the population level prior to invitation to the STHLM3 trial, meaning that the randomized intervention was “invitation to undergo a single prostate cancer screen using PSA and Stockholm3”. The randomization ratio was 70%, and it was chosen to ensure an invited group size to guarantee enough men enrolling in the STHLM3 to ensure statistical power for the primary analysis published in Grönberg et al. 2015^6^ comparing test characteristics of PSA vs. the Stockholm3 test (see *Predictive probability of success* Section for more details).

Men were randomly invited based on their date of birth between May 2012 and November 2014. Randomization time (invitation date, which we define as time 0 of the follow-up) corresponds to when letters were sent out inviting them to participate in the screening study. Non-invited men were assigned an invitation date (which we denote as “pseudo-invitation date”) that matched the STHLM3 invitations. The pseudo-invitation date will be considered as time 0 for the non-invited men.

All randomized participants are monitored through national or regional registries. Follow-up information on baseline characteristics and the outcomes will require data collection from various registries, including Statistics Sweden, the National Board of Health and Welfare, the National Prostate Cancer Register, and the Stockholm Prostate Cancer Diagnostics Register^9,10^.

#### Predictive probability of success

The STHLM3 trial’s sample size was determined to guarantee sufficient power for the primary analysis (published in 2015 in Grönberg et al., Lancet Oncology^6^). Therefore, we conducted an additional power analysis for the primary outcome of STHLM3-F/U using data collected at an interim monitoring after 6.5 years post-initiation of the STHLM3 trial (i.e. with a maximum of 6.5 years of follow-up)^11^. The power is a function of the number of events of the primary endpoint (prostate cancer mortality) and increases with follow-up time as the number events increase. We estimated the Bayesian predictive probability of success^12^ (PPoS) to guide the timing of the primary analysis of STHLM3-F/U, i.e. the time point when a sufficiently high number of prostate cancer deaths has occurred during follow-up to generate reasonably high statistical power for detecting a significant difference regarding the primary outcome of the trial^13^.

Success with respect to the primary outcome of the trial is evaluated by the p-value for the two-sided alternative hypothesis *H*_1_ : *RR*_*PCM*_ ≠ 1 versus the null *H*_0_ : *RR*_*PCM*_ = 1 calculated as 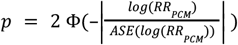, where *RR*_*PCM*_ is the risk ratio for prostate cancer mortality at the end of the follow-up time period, *ASE*(*log*(*RR*_*PCM*_)) is the asymptotic standard error for the *log*(*RR*_*PCM*_) and Φ(·) is the standard normal cumulative distribution function (more details in Section 3.3). The alpha level used for the interim analysis was 2%^11^. Assuming an information fraction of ≈30%, the final analysis will be done at an alpha level of 3.5% to ensure that the overall type I error rate, accounting for both interim and final analyses, remains at 0.05^14^.

Results from the interim analysis and the PPoS analysis were utilized to inform the selection of a specific time point for the final analysis, since the time point for the final comparison between the two invitation arms was only specified in the original STHLM3 study protocol to be between 10 and 15 years after study initiation^6^.

At the interim analysis, the crude cumulative risk of prostate cancer mortality (PCM) was compared between the two randomized groups at 6.5 years after invitation, and the difference in PCM risk was summarized by a risk ratio (RR) with the non-invited group as the referent. At the interim analysis 6.5 years after the initiation of the STHLM3 trial, the significance level α(6.5) was set to 0.02. The RR for PCM was 0.66 (98% CI: 0.41, 1.06), p=0.04, which favoured the invited group but did not reach alpha adjusted formal statistical significance.

We predicted the probability of observing a statistically significant RR at final analysis based on data gathered until 2018-12-31 (i.e. the data used in the 6.5 years interim analysis). Specifically, we estimated the PPoS by simulating scenarios of the final analysis, with follow-up until the end of 2022, 2023, and so on until 2027.

Since the PPoS with final analysis at any time between 2022 and 2027 was deemed sufficiently high and robust across the modeling strategies evaluated, the decision to compare PCM risk using follow-up until 2023-12-31 was made by the statisticians and principal investigators of the STHLM3 trial. The PPoS at the final analysis with data until 2023-12-31 is projected to reach 80%, ranging from 80 to 95% based on the modeling strategy used in the analysis^13^ .

For a comprehensive description of the PPoS calculation, readers are referred to https://doi.org/10.48550/arXiv.2412.15899.

## 3. Statistical analysis

### 3.1 Descriptive analyses

#### Trial flowchart

We will summarize the flow of study participants in a CONSORT diagram (Figure 2), including the number of eligible and randomized men, as well as the number of men undergoing the intervention. Further elements may also be included in the diagram.

#### Baseline characteristics

Descriptive characteristics will be reported to describe the population, presenting characteristics by randomisation group. Continuous variables will be summarized with mean and/or median and interquartile range, while categorical variables will be presented as counts and percentages. Variables of interest will include age, region of birth, civil status, educational level, income, prostate cancer family history, Charlson comorbidity index (CCI), and previous PSA testing or biopsy at invitation (baseline).

### 3.2 Analysis strategies

#### Effectiveness of the screening invitation

The main analyses will be conducted according to the intention-to-screen (ITS) principle, i.e. on all men included in the STHLM3-F/U cohort (i.e. on all men who were randomized to be invited or not invited to STHLM3) regardless of whether the invited men participated in STHLM3 or not. The ITS analysis will therefore compare the two randomized groups, i.e. men invited to the screening intervention and men not invited. This analysis targets the *effectiveness* of the screening invitation. All study outcomes, primary and secondary (see below), will be analyzed according to this principle.

#### Efficacy of the STHLM3 screening procedure

A secondary analysis will target the *efficacy* of the STHLM3 screening procedure. The aim is to estimate the effect in participants of the one-off screening using PSA and Stockholm3 test in combination, i.e. to compare the prostate cancer mortality risk (primary outcome; see below) in those men who received the STHLM3 invitation and chose to participate with the risk in those men randomly assigned to the non-invited group who would have participated if they had been invited.

First, we will use the method proposed by Cuzick et al^15^, as done in other prostate cancer screening trials (ERSPC^16^, CAP^17,18^), to adjust for non-compliance to the screening procedure. We will then use regression standardization as an alternative statistical method that targets the same quantity. Briefly, we will fit separate flexible parametric survival models for STHLM3 participants and non-invited men including age, region of birth (Nordic country vs non Nordic country), civil status, educational level, income, family history of prostate cancer, Charlson comorbidity index (CCI ≥ 1 vs CCI = 0), and previous PSA or biopsy exposure (yes vs no) at invitation as covariates. We will then standardize the model-based predicted prostate cancer mortality risks (in the presence of death due to other causes as a competing event) to the empirical distribution of the covariates in those men who were invited and participated in STHLM3.

The validity of these two efficacy analyses depends on the assumptions underlying the two statistical methods described above. As a benchmark, we will analyze the efficacy of the STHLM3 screening procedure on all-cause mortality. Since there is no reason to expect long-term differences in terms of all-cause mortality between participants and the counterfactual group of non-invited men who would have participated if they had been invited, any meaningful difference resulting from the analyses would indicate that the assumptions may not be (fully) met.

### 3.3 Primary outcome

Prostate cancer mortality, a time-to-event endpoint, will be considered as the primary outcome. The prostate cancer mortality risk at the end of follow-up will be calculated from the cumulative incidence functions (CIF) using the nonparametric Aalen-Johansen estimator, taking into account the presence of other-cause mortality as a competing event. Risk ratios (*RR*) will be calculated to compare the invitation groups at an alpha level of 3.5% against the null hypothesis *H*_0_ : *RR*_*PCM*_ = 1 . A two-sided alternative hypothesis will be adopted because the possibility that a screening invitation might increase the risk of prostate cancer mortality cannot be excluded a priori. For calculating p-values and confidence intervals, the asymptotic standard error derived from delta method for the log risk ratio (ASE(log(*RR* _*PCM*_))) will be used. The non-invited group will be the reference group for the *RR*.

Date of invitation will be used as time 0. Follow-up time will be calculated as the time between the date of invitation and the date of the earliest event between death (from prostate cancer or from other causes, respectively), emigration outside Sweden, or end of follow-up (2023-12-31, administrative censoring).

Prostate cancer deaths will be identified using the National Cause of Death Register^19^, and all deaths with C61 as “underlying cause of death according to ICD-10” will be considered prostate cancer deaths^20^.

### 3.4 Secondary outcomes

For secondary outcomes, point estimates and 95% confidence intervals in the two randomized groups will be presented, and may be compared using ratios.

#### All-cause mortality

All-cause mortality will be evaluated at the end of follow-up using non-parametric cumulative incidence functions, comparing groups using risk ratios. Date of invitation will be used as time 0. Follow-up time will be calculated as the time between the date of invitation and the first event between emigration outside Sweden, death or end of follow-up (administrative censoring).

#### Prostate cancer incidence

Incidence of prostate cancer will be evaluated at the end of follow-up, overall (Gleason Score ≥6), and differentiating between Gleason Score 6 and Gleason Score ≥7. Incidence analyses will also be conducted based on Gleason Score at diagnosis, grouping cases into categories of GS 3+3, 3+4, 4+3, and ≥8. Incidence analyses will use the same method as for prostate cancer mortality, comparing groups with risk ratios. Death from any causes will be considered as a competing event.

Date of invitation will be used as time 0. Follow-up time will be calculated as the time between the date of invitation and the first event between prostate cancer diagnosis, emigration outside Sweden, death or end of follow-up (administrative censoring).

Non-missing Gleason Score information is required for the incidence analysis grouped by Gleason Score. Missing values will be imputed using Multivariate Imputation by Chained Equations (MICE)^21^ with predictive mean matching (PMM) and a donor pool of k=5. Specifically, missing data for individual primary and secondary Gleason patterns (GS1 and GS2) will be imputed using age at invitation, date of diagnosis, and PSA at diagnosis as predictors. We will generate 1000 datasets with imputed outcome data, which will be analyzed separately. We will then use Rubin’s rules to pool risks (and standard errors) of prostate cancer diagnosis in each group.

#### PSA and Biopsy patterns

The fraction of men undergoing at least one PSA test and biopsy will be evaluated, as well as the cumulative number of tests performed during the follow-up. The mean number of PSA tests and biopsies per man over the follow-up period will also be estimated.

Only diagnostic PSA tests and biopsies will be considered, meaning tests conducted on men who are free of a prostate cancer diagnosis, excluding those tests taken to monitor diagnosed patients. All PSA tests performed before the date of prostate cancer diagnosis will be included. To account for potential delays in biopsy reporting, biopsies conducted up to 30 days after diagnosis will also be included. This 30-day time allowance permits sufficient reporting time, as recommended by clinician advisors, ensuring the inclusion of actual diagnostic biopsies.

Date of invitation will be used as time 0. PSA tests and biopsies related to the intervention will be included in the analysis. Men will be followed until the first event between prostate cancer diagnosis (+30 days in the biopsy analysis), emigration, death, or end of follow-up (administrative censoring).

To estimate the mean number of tests per man over the follow-up period, we will use the marginal Ghosh and Lin non-parametric method accounting for competing events^22^, that will consist of mortality and prostate cancer diagnosis.

#### Stockholm3 test

Since 2018 the use of the Stockholm3 test has been introduced in clinics across the Stockholm region. We may, depending on the availability of the data, report the use of the Stockholm3 test over the follow-up period. Proportion of men tested with the Stockholm3 test in each randomized group will be calculated, overall and stratified by year. Similar analysis as for the PSA test may be performed (see preceding section).

### 3.5 Additional analysis – different measures

Since STHLM3 is a one-time screen (i.e. the men were invited to screening only once), it is likely that any difference between the groups (i.e. men invited to STHLM3 vs. not) with respect to prostate cancer mortality will increase in the years following the intervention and then at some point start to decrease again. The reason why this is likely is that prevalent cases (cases present in the population at time of invitation, but yet undetected) will to a higher degree be detected in the intervention arm (i.e. among men invited to screening), however we do not expect large differences between the groups with respect to incident development of new cases *after* the STHLM3 intervention. Eventually, the prostate cancer mortality from the incident cases detected after the STHLM3 intervention may start to dominate the total number of men who die due to prostate cancer, meaning that the difference in mortality between the groups may start to decrease again. Therefore, in addition to analyzing the risk ratio at the end of follow-up, we will also compute the risk ratio (and 95% confidence intervals) at additional time points (Section 3.7). Additionally, we may also compare groups using restricted mean time lost to evaluate different estimands of the effect of the intervention (randomization to receive an invitation to the STHLM3 screening trial vs. not). For the same reason, cause-specific hazard ratios from Cox models will be estimated.

### 3.6 Subgroup analysis

We will perform subgroup analyses by stratifying men based on age groups at the time of invitation (in 5-year strata) and educational level (low-mid-high) as a measure of socioeconomic status. Income may also be used for the same purpose as educational level.

#### STHLM3 participants

We may perform a subgroup analysis restricted to STHLM3 participants, to investigate potential heterogeneity within this group. Since the STHLM3 trial had a paired design, men could be referred to further diagnostic workup (biopsy) based on elevated PSA test (≥3 ng/mL), elevated Stockholm3 test (≥10%), or both. The STHLM3 trial participants can be categorised based on their PSA and Stockholm3 value at screening into 4 risk groups: (+,+), (+,-), (-,+), (-,-). We may investigate mortality stratified over these four groups. We may also investigate mortality among participants based solely on PSA value at STHLM3 screening, for example by stratifying men into groups: PSA ≤ 1, between 1 and 2, between 2 and 3, and PSA ≥3 ng/mL.

### 3.7 Sensitivity analysis

#### Analysis without multiple imputation

We will repeat the incidence analysis among different Gleason Score groups without imputation of missingness on Gleason Score.

#### Analysis at different time points

The main analysis will be performed at the end of available follow-up. We will repeat the analysis on mortality and incidence at different time points of interest over the follow-up period, by extracting risks from the cumulative incidence functions at specific time points (eg: 4, 6, 8, 10 years of follow-up).

#### STHLM3-MRI trial

The STHLM3-MRI study (NCT03377881)^23^ has been ongoing in the Stockholm region since 2018^24–26^. In the STHLM3-MRI study, the Stockholm3 test was used together with the PSA test. We anticipate an increase in prostate cancer incidence during the years in which the trial was conducted due to the organized screening among the STHLM3-MRI participants.

We can retrieve information on the men invited to participate in the STHLM3-MRI trial and those who participated in the trial. We will conduct an analysis of mortality and incidence, excluding men invited to the STHLM3-MRI trial. Since these men were invited to participate in the STHLM3-MRI trial conditional on *not* having participated in the STHLM3 trial, their exclusion will break randomization in the STHLM-F/U trial. We may account for this using statistical methods such as inverse probability of treatment weighting or regression standardisation.

## 4. Discussion

This study aims to address the knowledge gap regarding the long-term effect of screening with the Stockholm3 test, a diagnostic tool designed to improve upon the limitations of the traditional PSA test. It will provide a novel analysis of mortality data from men screened for prostate cancer using both PSA and Stockholm3 (a prediction model incorporating novel biomarkers, clinical variables, and a genetic risk score).

The design of the STHLM3 trial included two nested interventions: first a *randomized invitation* to the STHLM3 trial and then a *paired intervention* among those men who were invited to and participated in STHLM3 to assess the operating characteristics of the Stockholm3 test relative to those of the PSA test. The design with the two nested interventions allows us to use the randomized groups to evaluate the effects of the invitation. Understanding whether a systematic screening intervention, employing an approach to screening using both PSA and the Stockholm3 test, has any effect on mortality can help inform decisions on prostate cancer screening policies.

## Data Availability

All data used in this work are included in the manuscript. Since this manuscript describes a protocol, no outcome data has been collected yet.

## 5. Ethics

The study protocol was approved by Stockholm regional ethics committee (permits 2012/572-31/1 and 2012/438-31/3). Written informed consent was obtained from all participants in the STHLM3 intervention.

